# Prognostic value of C-reactive protein in patients with COVID-19

**DOI:** 10.1101/2020.03.21.20040360

**Authors:** Xiaomin Luo, Wei Zhou, Xiaojie Yan, Tangxi Guo, Benchao Wang, Hongxia Xia, Lu Ye, Jun Xiong, Zongping Jiang, Yu Liu, Bicheng Zhang, Weize Yang

**Author notes:** Correspondence to: Prof Xiaomin Luo MD, Department of Emergency, Eastern Campus, Renmin Hospital of Wuhan University, Wuhan 430060, China., Prof Bicheng Zhang MD, Cancer center, Renmin Hospital of Wuhan University, Wuhan 430060, China., Prof Weize Yang MD, Department of Emergency, Eastern Campus, Renmin Hospital of Wuhan University, Wuhan 430060, China. Xiaomin Luo, Wei Zhou, Xiaojie Yan, and Tangxi Guo contributed equally to this manuscript. Xiaomin Luo, Bicheng Zhang, and Weize Yang are joint corresponding authors. **Main points of this manuscript:** In patients with COVID-19, admission CRP correlated with disease severity and tended to be a good predictor of adverse outcome. Patients with markedly elevated admission CRP should be provided more attention and strengthened treatment.

## Abstract

**Background:** Elevated serum C-reactive protein (CRP) level was observed in most patients with COVID-19.

**Methods:** Data of COVID-19 patients with clinical outcome in a designated hospital in Wuhan, China, were retrospectively collected and analyzed from Jan 30 to Feb 20, 2020. The prognostic value of admission CRP was evaluated in patients with COVID-19.

**Results:** Out of 298 patients enrolled, 84 died and 214 recovered. Most non-survivors tended to be males, old aged, or with chronic diseases. Compared to survivors, non-survivors showed significantly elevated white blood cell and neutrophil count, neutrophil to lymphocyte ratio (NLR), systemic immune-inflammation index (SII, defined by platelet count multiply by NLR), CRP, procalcitonin, and D-dimer, and decreased red blood cell, lymphocyte, and platelet count. Age, neutrophil count, platelet count, and CRP were identified as independent predictors of adverse outcome. The area under the receiver operating characteristic (ROC) curve (AUC) of CRP (0.896) was significantly higher than that of age (0.833), neutrophil count (0.820), and platelet count (0.678) in outcome prediction (all p<0.05). With a cut-off value of 41.4, CRP exhibited sensitivity 90.5%, specificity 77.6%, positive predictive value 61.3%, and negative predictive value 95.4%. Subgroup analysis revealed that CRP remained robust accuracy in adverse outcome prediction in patients with different disease severity (AUC 0.832, z=10.23, p<0.001; AUC 0.989, z=44.04, p<0.001). CRP was also an independent discriminator of severe/critical illness on admission (AUC 0.783, z=10.69, p<0.001).

**Conclusions:** In patients with COVID-19, admission CRP correlated with disease severity and tended to be a good predictor of adverse outcome.

## Introduction

Since the first cases of novel coronavirus pneumonia, latterly named COVID-19 by World Health Organization (WHO), were reported in Wuhan in December 2019, SARS-Cov-2 has infected 81174 patients and caused 3242 deaths in China.^1-4^ This newly discovered coronavirus was named SARS-Cov-2 by WHO due to its similarity in gene sequence to severe acute respiratory syndrome coronavirus (SARS-CoV).^2^ On 30 Jan 2020, WHO declared COVID-19 epidemic a public health emergency of international concern.^2^ In order to control SARS-Cov-2 epidemic timely, multiple active measures have been taken in Wuhan, the worst epidemic area in China, including centralized community quarantine, recruiting designated hospitals, establishing module hospitals, and so on. SARS-CoV-2 epidemic has been preliminary controlled in Wuhan, China at current with the strengthened support from central and local governments. However, confirmed cases worldwide have exceeded 200,000 by 19 Mar 2020. Many other countries such as Italy, Iran, Spain, and United States of America, however, are showing an ongoing outbreak,^4^ which will inevitably bring about shortage of medical resources. According to the report by Yang et al., the 28-day mortality was 61.5% for a group of critically ill COVID-19 patients.^5^ Many patients with mild symptoms presented a sudden progressive to severe or critical illness.^6^ Hence, to identify simple and efficient predictor is vital for us to provide strengthened attention and treatment to the targeted patients and thus to reduce the mortality of COVID-19.

Similar to SARS, critical patients with COVID-19 presented higher plasma levels of cytokines, suggesting the involvement of inflammatory storm in the pathogenesis of disease progression.^7^ C-reactive protein (CRP), a routinely measured inflammatory marker, was increased in most patients with COVID-19 and associated with disease severity.^7-9^ In a Swedish multicenter study, CRP was suggested to be a simple, early marker for prognosis in intensive care unit (ICU) admissions for sepsis. An admission CRP level above 100 mg/L was found to be associated with increased ICU and 30-day mortality.^10^ To date, the prognostic value of CRP has not been tested in patients with COVID-19. In this retrospective study, we aimed to evaluate the potential of CRP in outcome prediction of patients with COVID-19.

## Methods

### Study design and participants

This retrospective, single-center study was conducted at Eastern Campus of Renmin Hospital of Wuhan University from Jan 30 to Feb 20, 2020. Eastern Campus was requisitioned as a designated hospital for COVID-19 on Jan 25, 2020 and began to admit patients on Jan 30, 2020 after being remodeled. All adult patients who had clinical outcome (died or recovered) till Feb 20, 2020 were recruited in this study. Patients without CRP detection on admission were excluded. The confirmation of patients with COVID-19 was according to guidance issued by NHC.^11^ This study was approved by the Ethics Committee of Renmin Hospital of Wuhan University (approval number: WDRM 2020-K094). Written consent was not required for the retrospective character of this study.

### Data collection

Information of each patient was obtained mainly through screening Electronic Health Records and Laboratory Information Management System provided by DHC Software Co., Ltd (Beijing, China). Nursing records were also reviewed if necessary. Epidemiological information such as gender, age, chronic diseases, history of smoking and drinking, were reviewed and assessed, as well as days from illness onset to hospitalization and disease severity on admission. Results of some laboratory test on admission were collected and evaluated, including complete blood cell count, neutrophil to lymphocyte ratio (NLR), systemic inflammatory index (SII, defined by platelet count multiply by NLR), CRP, procalcitonin, and D-dimer. Disease severity (ordinary, severe or critical) was determined according to the guidance issued by NHC.^11^ The primary endpoint was clinical outcome (death or recovery), and the secondary endpoint was disease severity on admission.

### Statistical analysis

Categorical variables were presented as number (%) and compared by Chi-squared test. Normal distribution of continuous variables was evaluated by the Kolmogorov-Smirnov test. Normally or non-normally distributed continuous parameters were described as mean (standard deviation, SD) or as median (interquartile range, IQR), and were compared by independent t test or Mann-Whitney test, respectively. Multivariate logistical regression was conducted to identify independent risk factors of study endpoints. The accuracy of each independent predictor was determined by each area under the receiver operating characteristic (ROC) curve (AUC). The Hosmer and Lemeshow test for goodness-of-fit statistics was used to check model adequacy. The AUCs of independent predictors were compared by Hanley-McNeil test. All statistical analyses were performed using IBM SPSS Statistics 19.0 and Medcalc software 16.2. A two-sided test of α < 0.05 was considered statistical significance.

## Results

### Clinical characteristics of patients with COVID-19

By Feb 20, 2020, 359 patients had clinical outcome, among whom, 61 were excluded due to lack of CRP data on admission. Hence, 298 patients were finally enrolled in this study, including 141 cases of ordinary illness and 157 cases of severe or critical illness on admission. Eight four patients died and 214 recovered. There were 150 males and 148 females, with median age of 57(40-69) years. One hundred and thirty-five (45.2%) patients had chronic diseases. There were 86 cases of hypertension, 32 cases of cerebrovascular diseases, 45 cases of diabetes, 26 cases of coronary heart disease, 23 cases of chronic pulmonary disease, 16 cases of cirrhosis, and 12 cases of anemia. Most patients who died were males, aged over 60 years or with chronic diseases. Twenty-one (7.0%) patients had a history of smoking and 33 (11.1%) had a history of drinking. The median time from onset of symptoms to hospital admission was 9 (6-12) days. No significant difference existed in history of smoking or drinking, and days from illness onset to hospitalization between survivors and non-survivors. Non-survivors showed significantly higher white blood cell and neutrophil count, NLR, SII, CRP, procalcitonin, and D-dimer, and lower red blood cell, lymphocyte, and platelet count compared to survivors (Table 1).

**Table 1.** Epidemiological and laboratory findings of patients with COVID-19

|  | All patients (n=298) | Died (n=84) | Recovered (n=214) | *P* value |
| --- | --- | --- | --- | --- |
| Age, years | 57 (40-69) | 71 (64-80) | 51 (37-63) | 0.000 |
| Gender |  |  |  | 0.025 |
| Male | 150 (50.3%) | 51 (60.7%) | 99 (46.3%) |  |
| Female | 148 (49.7%) | 33 (39.3%) | 115 (53.7%) |  |
| Smoking | 21 (7.0%) | 8 (9.5%) | 13 (6.1%) | 0.295 |
| Drinking | 33 (11.1%) | 10 (11.9%) | 23 (10.7%) | 0.775 |
| With chronic diseases | 135 (45.2%) | 61 (72.6%) | 74 (34.6%) | 0.000 |
| Hypertension | 86 (28.9%) | 49 (58.3%) | 37 (17.3%) | 0.000 |
| Cerebrovascular diseases | 32 (10.7%) | 17 (20.2%) | 15 (7.0%) | 0.001 |
| Coronary heart disease | 26 (8.7%) | 13 (15.5%) | 13 (6.1%) | 0.010 |
| Diabetes | 45 (15.1%) | 18 (21.4%) | 27 (12.6%) | 0.056 |
| Chronic pulmonary disease | 23 (7.7%) | 13 (15.5%) | 10 (4.7%) | 0.002 |
| Cirrhosis | 16 (5.4%) | 5 (6.0%) | 11 (5.1%) | 0.782 |
| Chronic kidney disease | 5 (1.7%) | 2 (2.4%) | 3 (1.7%) | 0.928 |
| Anemia | 12 (4.0%) | 5 (6.0%) | 7 (3.3%) | 0.464 |
| Days from onset to admission | 9 (6-12) | 9 (7-14) | 8 (6-11) | 0.187 |
| Disease severity |  |  |  | 0.000 |
| Ordinary illness | 141 (47.3%) | 4 (4.8%) | 137 (64.0%) |  |
| Severe or critical illness | 157 (52.7%) | 80 (95.2%) | 77 (36.0%) |  |
| Laboratory findings |  |  |  |  |
| White blood cell count, $\times 10^9/L$ | 5.63 (4.12-7.47) | 8.58 (5.26-12.70%) | 5.19 (3.98-6.48) | 0.000 |
| Neutrophil count, $\times 10^9/L$ | 3.87 (2.68-5.78) | 6.92 (4.33-10.79) | 3.20 (2.53-4.56) | 0.000 |
| Lymphocyte, $\times 10^9/L$ | 1.03 (0.78-1.44) | 0.83 (0.63-1.09) | 1.04 (0.83-1.50) | 0.000 |
| Red blood cell count, $\times 10^{12}/L$ | 4.14 (0.64) | 3.99 (0.71) | 4.20 (0.61) | 0.013 |
| Hemoglobin, g/L | 127.3 (18.0) | 125.5 (20.0) | 128.0 (17.2) | 0.280 |
| Platelet count, $\times 10^9/L$ | 190 (143-244) | 154 (111-213) | 205 (151-252) | 0.000 |
| Neutrophil to lymphocyte ratio | 3.72 (2.42-7.25) | 8.17 (6.15-10.90) | 2.96 (2.13-4.61) | 0.000 |
| systemic inflammatory index | 710 (435-1246) | 1223 (743-1847) | 563 (390-882) | 0.000 |
| C-reactive protein, mg/L | 25.5 (5.0-80.1) | 100.0 (60.7-179.4) | 9.65 (5.0-37.9) | 0.000 |
| D-Dimer, mg/L | 0.75 (0.37-3.42) | 4.59 (0.95-17.14) | 0.50 (0.29-1.10) | 0.000 |
| Procalcitonin, ng/mL | 0.057 (0.034-0.137) | 0.228 (0.119-0.991) | 0.043 (0.027-0.065) | 0.000 |

Red blood cell count and hemoglobin were presented as mean (SD) and compared with independent t test between non-survivors and survivors. Other continuous variables were presented as median (IQR) and compared with Mann-Whitney test. Chi-squared test was conducted to compare categorical variables between two groups.

### Independent predictors for adverse clinical outcome

The independent predictors were identified from risk factors relating to adverse clinical outcome by logistic regression model. As indicated in table 2, age, neutrophil count, platelet count, and CRP were found to be independent predictors of adverse outcome. Hosmer and Lemeshow test showed a good model adequacy *(χ*^2^=6.64, p=0.576). To determine and compare the accuracy of these factors in adverse outcome prediction, ROC curve analysis was performed and the difference in AUCs was tested. The AUCs of age, neutrophil count, platelet count, and CRP were 0.833 (95% CI: 0.782–0.884) (z=12.77, p<0.001), 0.820 (95% CI: 0.761–0.880) (z=10.52, p<0.001), 0.678 (95% CI: 0.609–0.747) (z=5.06, p<0.001), and 0.896 (95% CI: 0.857–0.935) (z=19.76, p<0.001), respectively (Table 3, Figure 1). The AUC of CRP was significantly higher than that of age (z=2.05, p=0.041), neutrophil count (z = 2.09, p =0.028), and platelet count (z = 5.65, p <0.001) for adverse outcome prediction. With a cut-off value of 41.4, CRP exhibited sensitivity 90.5%, specificity 77.6%, positive predictive value (PPV) 61.3%, and negative predictive value (NPV) 95.4%.

**Table 2.** Independent predictors of adverse outcome in patients with COVID-19

|  | B | SD | Odds ratio (95% CI) | P value |
| --- | --- | --- | --- | --- |
| Age | -0.072 | 0.017 | 0.930 (0.900-0.962) | 0.000 |
| Neutrophil count | -0.533 | 0.111 | 0.587 (0.472-0.730) | 0.000 |
| Platelet count | 0.015 | 0.004 | 1.015 (1.007-1.024) | 0.000 |
| C-reactive protein | -0.020 | 0.005 | 0.980 (0.971-0.990) | 0.000 |

B: non-standard coefficient; CI: confidence intervals

**Table 3.** Independent predictors of adverse outcome in subgroup analysis according to disease severity

|  |  | B | SD | Odds ratio (95% CI) | P value |
| --- | --- | --- | --- | --- | --- |
| Severe/critical | Age | -0.059 | 0.019 | 0.943 (0.909-0.978) | 0.002 |
|  | Neutrophil count | -0.557 | 0.126 | 0.573 (0.448-0.733) | 0.000 |
|  | C-reactive protein | -0.017 | 0.005 | 0.983 (0.974-0.992) | 0.000 |
|  | Platelet count | 0.017 | 0.005 | 1.017 (1.008-1.027) | 0.000 |
| Ordinary | C-reactive protein | -0.053 | 0.019 | 0.948 (0.913-0.985) | 0.006 |

B: non-standard coefficient; CI: confidence intervals

**Figure 1.**
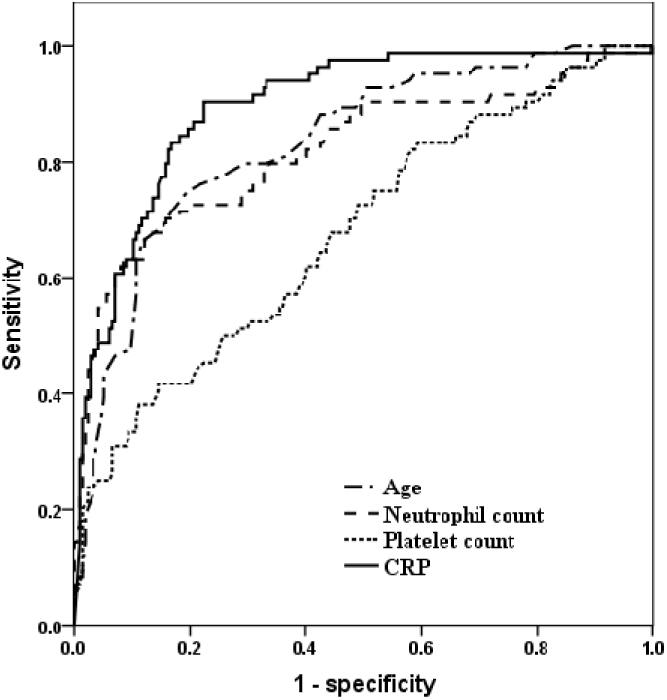
Receiver operating characteristic (ROC) curves of age, neutrophil count, platelet count, and C-reactive protein for adverse outcome prediction. The area under the curve (AUC) of age, neutrophil count, platelet count, and C-reactive protein for predicting adverse outcome was 0.833, 0.820, 0.678, and 0.896, respectively. The AUC of C-reactive protein was significantly higher than that of age (z=2.05, p=0.041), neutrophil count (z = 2.09, p =0.028), and platelet count (z = 5.65, p <0.001) for adverse outcome prediction.

Subgroup analysis in adverse outcome prediction was performed for patients according to different disease severity on admission. In this case, age, neutrophil count, platelet count, and CRP were still identified as independent risk factors of adverse outcome in patients with severe or critical illness (table 3). Hosmer and Lemeshow test showed a good model adequacy (*χ*^2^=4.99, p=0.759). The AUCs of age, neutrophil count, platelet count, and CRP for adverse outcome prediction were 0.726 (95% CI: 0.646 - 0.806) (z=5.53, p<0.001), 0.787 (95% CI: 0.716 - 0.859) (z=7.84, p<0.001), 0.697 (95% CI: 0.618 - 0.767) (z=4.76, p<0.001), and 0.832 (95% CI: 0.768 - 0.896) (z=10.23, p<0.001), respectively. The AUC of CRP was comparable to that of neutrophil count (z=0.98, p = 0.326), but significantly higher than those of age (z=2.06, p = 0.039) and platelet count (z=2.70, p = 0.007). With a cut-off value of 56.3, CPR showed sensitivity 81.3%, specificity 71.4%, PPV 74.7%, and NPV 78.6%. For patients with ordinary illness, CRP was identified as an only independent predictor of adverse outcome (table 3), with AUC 0.989 (95% CI: 0.967-1.000) (z=44.04, p<0.001). With a cut-off value 0f 80.9, CRP showed sensitivity 100.0%, specificity 95.6%, PPV 40.0%, and NPV 100.0%.

### Independent discriminators of severe/critical illness on admission

Among 150 male patients, 91 were diagnosed as severe/critical illness, while 59 were diagnosed as ordinary illness at admission. Patients with severe/critical illness tended to present a larger percent of male gender compared to patients with ordinary illness (58% vs. 41.8%, *χ*^2^=7.72, p=0.005). A much lower percent of chronic diseases was observed in patients with ordinary illness (29.1% vs. 59.9%, *χ*^2^ =28.43, p<0.001). Compared to patients with ordinary illness, patients with severe/critical illness showed older age (median 67 (57-75) years vs. 42 (32-57) years, p<0.001) and more duration from illness onset to admission (median 10 (7-13) days vs. 8 (5-10) days, p<0.001). Raised white blood cell count (median 6.63 (4.52-9.27) × 10^9^/L vs. 5.12 (3.92-6.16) × 10^9^/L, p<0.001) and neutrophil count (median 4.99 (3.18-8.14) × 10^9^/L vs. 3.10 (2.50-4.21) × 10^9^/L, p<0.001), and reduced red blood cell count (4.00 (0.68) × 10^12^/L vs. 4.30 (0.57) × 10^12^/L, p<0.001), lymphocyte count (median 0.89 (0.71-1.17) × 10^9^/L vs. 1.06 (0.86-1.60)× 10^9^/L, p<0.001), platelet count (median 177 (134-240) × 10^9^/L vs. 205 (151-250) × 10^9^/L, p=0.043) and hemoglobin level (124 (19) g/L vs. 131 (17) g/L, p=0.001) were observed in patients with severe or critical illness compared to those with ordinary illness. Patients with severe or critical illness tended to exhibit elevated NLR (median 6.26 (3.49-9.12) vs. 2.58 (1.86-3.70)), SII (median 1053 (581-1612) vs. 488 (358-779)), CRP (median 60.8 (21.0-110.6) mg/L vs. 7.7 (5.0-29.3) mg/L), procalcitonin (median 0.093 (0.047-0.266) ng/mL vs. 0.040 (0.021-0.062) ng/mL), and D-dimer (median 1.21 (0.52-6.86) mg/L vs. 0.40 (0.24-0.90) mg/L) (all p<0.001). As indicated in table 4, age, neutrophil count, and CRP were verified to be independent discriminators of disease severity on admission by multivariate logistic regression analysis. Hosmer and Lemeshow test showed a good model adequacy (χ^2^=8.31, p=0.404). The AUCs of these discriminators were 0.828 (95% CI: 0.781-0.875) (z=13.69, p<0.001), 0.729 (95% CI: 0.671-0.786) (z=7.79, p<0.001), and 0.783 (95% CI: 0.731-0.835) (z=10.69, p<0.001), respectively. The AUC of CRP was comparable to that of age (z=0.145, p=0.147) and neutrophil count (z=1.54, p=0.124) (Figure 2). With a cut-off value of 41.3, CRP exhibited sensitivity 65.0%, specificity 83.7%, PPV 81.6%, and NPV 68.2%.

**Table 4.** Independent discriminators of disease severity on admission

|  | B | SD | Odds ratio (95% CI) | P value |
| --- | --- | --- | --- | --- |
| Age | 0.045 | 0.011 | 1.047 (1.024-1.070) | 0.000 |
| Neutrophil count | 0.252 | 0.072 | 1.286 (1.117-1.481) | 0.000 |
| C-reactive protein | 0.009 | 0.004 | 1.009 (1.002-1.017) | 0.010 |

B: non-standard coefficient; CI: confidence intervals

**Figure 2.**
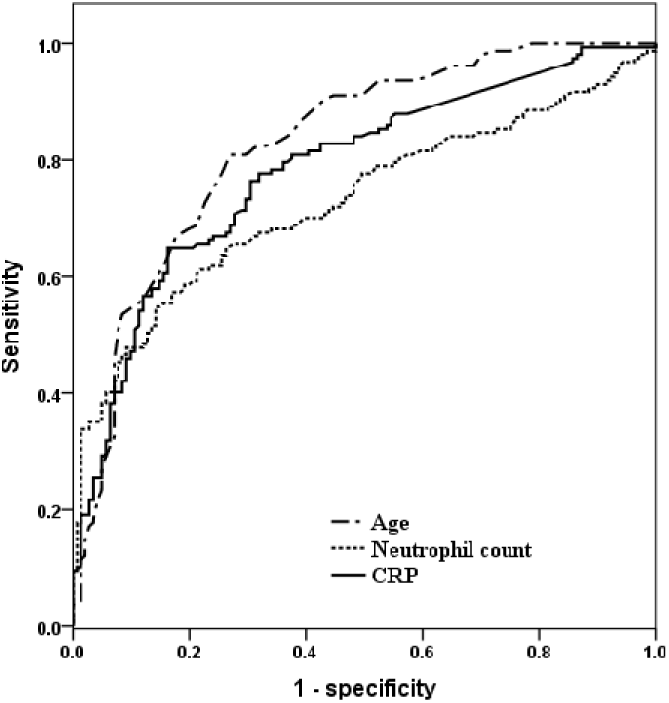
ROC curves of age, neutrophil count, and C-reactive protein for discriminating disease severity on admission. The area under the curve (AUC) of age, neutrophil count, and C-reactive protein was 0.828, 0.729, and 0.783, respectively. The AUC of C-reactive protein was comparable to that of age (z=0.145, p=0.147) and neutrophil count (z=1.54, p=0.124)

## Discussion

In this retrospective study, age, neutrophil count, platelet count, and CRP were verified to be independent outcome predictors in patients with COVID-19, while age, neutrophil count, and CRP were identified to be independent discriminators of disease severity on admission. Some important biomarkers of infection or critical illness including NLR, SII, procalcitonin, and D-dimer, were found to be associated with clinical outcome and disease severity. However, none of them were identified as independent predictors. The findings of this study suggested that CRP performed better than other three parameters in predicting adverse outcome in patients with COVID-19. Besides, admission serum CRP level was identified as a moderate discriminator of disease severity. To our knowledge, this study presented the first report about the prognostic value of CRP in patients with COVID-19.

The pathological mechanism of COVID-19 has not been fully uncovered. In this study, the median age of non-survivors was 71 (64-80) years, significantly higher than 57 (40-69) years in survivors. Further, age was an independent predictor of adverse outcome and discriminator of severe/critical illness, which suggested that old people are more vulnerable to SARS-CoV-2 and more likely to develop severe/critical disease.^5,6,8^ Elevated neutrophil count was observed in patients with severe illness compared to those with non-severe illness.^7^ In this study, admission neutrophil count was a moderate predictor of clinical outcome and disease severity. These results implied that factors contributing to raised neutrophil count, such as secondary infection, excessive inflammatory stress or glucocorticoids use, might exacerbate disease progression in patients with COVID-19. Reduction in peripheral lymphocyte count was commonly observed in patients with COVID-19, which was considered a possible critical factor associated with disease severity and mortality.^12,13^ Reduced CD4 and CD8 T cell count accompanied by their over-activation might contribute to the impaired immunity and disease progression in patients with COVID-19.^12^ In this study, significant discrepancy was observed in lymphocyte count between patients with different outcome or disease severity. NLR, an inflammatory index defined by neutrophil count divided by lymphocyte count, was found to be associated with sepsis and multiple organ damage.^14^ Recently, a prospective study suggested NLR an early predictor of COVID-19 progressive to severe illness, however, the power of NLR in outcome prediction was not determined due to no follow-up of final outcome.^15^ In this retrospective study, raised NLR was found in non-survivors compared to survivors and associated with disease severity on admission. However, results of multivariate logistical regression revealed that neither lymphocyte count nor NLR was an independent predictor of adverse outcome or discriminator of severe/critical illness.

As one of the most distinctive acute phase reactants, CRP can increase rapidly after the onset of inflammation, cell damage or tissue injury. Pulmonary diseases with inflammatory features usually raise serum CRP level in response to inflammatory cytokines such as IL-6, IL-1 or TNF-α.^16,17^ Hence, markedly elevated serum CRP level in non-survivors or patients with severe/critical illness in this study indicated excessive inflammatory response, which was consistent with raised serum pro-inflammatory cytokines observed in COVID-19 patients.^18,19^ The roles of CRP in case of diseases may involve host defense and inflammation. In response to inflammatory onset, CRP binds to pathogens and promotes their elimination by phagocytic cells, functioning as the first line of innate host defense. Besides, CRP can exhibit anti-inflammatory effects by inhibiting neutrophils chemotaxis.^20^ However, by up-regulating expression of adhesion molecules, and pro-inflammatory IL-1, IL-6, IL-8, and TNF-α, CRP can also exert pro-inflammatory effects.^21^ Controversial results of serum CRP level were observed in patients with acute lung injury (ALI) or acute respiratory distress syndrome (ARDS).^22,23^ The association between high serum CRP level and a favorable outcome was found in adult patients with ALI/ARDS.^22^ In elderly patients with ALI, however, high serum CRP level correlated to higher mortality,^23^ which is consistent to our findings in patients with COVID-19. Direct attacks from SARS-CoV-2 and organ damage caused by excessive inflammatory response might be responsible for the pathogenesis of disease progression.^20^ Therefore, markedly elevated serum CRP level in patients with COVID-19 might be the indication of excessive inflammatory stress and contribute to severe/critical illness or even death. Nevertheless, the exact function of CRP in patients with COVID-19 remains unclear. Future researches should be focused on involvements of CRP in the pathogenesis of COVID-19.

There are some limitations in our study. First, aging, chronic diseases, and secondary infection in some cases might exert effects on the increased serum CRP level besides SARS-CoV-2 infection itself. Such superimposed effects, however, would better reflect the features of patients with severe COVID-19.

Second, some patients with critical illness were not admitted to intensive care unit as they should be due to shortage of resources, which undoubtedly had a negative impact on the outcome of those patients. Third, patients were not recruited consecutively due to exclusion of some cases without serum CRP detection on admission, which might bring about selective bias. Fourth, serum CRP level correlates with the degree of inflammatory response,^24^ therefore, changes in CRP over time might provide more information about disease prognosis. However, no continuous CRP values over time were included in this study since it was designed to explore the potential of admission CRP in outcome prediction.

In conclusion, our results suggested that admission serum CRP level performed well in discriminating disease severity and predicting adverse outcome in patients with COVID-19. Patients with markedly elevated admission CRP should be provided more attention and strengthened treatment. The findings from this single-center study need to be validated by multi-center research with larger samples.

## Data Availability

The data sets that support the findings of this study are available from the corresponding author on reasonable request.

## Acknowledgments

We thank all patients and their families involved in the study.

